# Serious Gaming and Eye-Tracking for the Screening, Monitoring, Diagnosis and Treatment of Neurodevelopmental Disorders in Children: A Systematic Literature Review

**DOI:** 10.1101/2025.09.16.25335869

**Authors:** Muhammad Farooq Shaikh, Ciara Higley, Cecilia Campanile, Rebecca Francis, Elyssa Panja, Silvia Santacaterina, Giacomo Pratesi, Davide Piaggio

## Abstract

Neurological development between the ages of 3 to 11 is crucial to the shaping of infrastructural capabilities like the executive functions that enable the child to achieve academically and socially. Such development can be hindered by neurodevelopmental disorders (NDDs) like attention deficit hyperactivity disorder (ADHD), Dyslexia, and Dysgraphia, which affect 5-10% of the world population of children. Although the importance of early screening is acknowledged, inadequacies such as access barriers, long waiting time, and excessive cost lead to late detection, even when potential issues are identified. This PRISMA-based systematic review examines the role of technology and serious games that may screen and treat NDDs in children early. The PubMed and Scopus databases were utilized, and research published between 2013, and February of 2025 was reviewed, where the age interval of the sampled children was between 3 and 11, and extended to 21 in relevant cases. Some of the tools reviewed are eye-tracking systems, machine learning models, mobile applications, and serious games. The quality of studies was assessed by the Mixed Methods Appraisal Tool (MMAT) and the results synthesized narratively. Out of 3,129 records, 37 studies were included according to the inclusion criteria. Findings indicated that although numerous technologies showed promise in recognizing and assisting children with NDDs, the majority had limited capabilities in scalability, longitudinal tracking, and practical application as the following was minimal, and the length of follow-up was low. In summary, the possibilities of using technology to better diagnose and intervene early are promising, although cost, training and implementation frameworks aligned with the NHS are critical barriers.

## 1. Introduction

Neurodevelopmental disorders are a group of disorders causing deficits in cognition and delayed brain development due to various causes, including dysregulation of mechanisms governing the development of the cerebral cortex (1). Currently, it is reported that 5-11% of children under 18 have been diagnosed with attention-deficit/hyperactivity disorder (ADHD), 3-10% with a specific learning disorder (SLD), and 0.63% with an intellectual disability. There is limited data available from low-income countries on neurodevelopmental disorders (2), which accounts for a discrepancy between the accurate prevalence of neurodevelopmental disorders (3, 4). The main neurodevelopmental disorders we are focusing on are dyslexia, dyscalculia, dysgraphia, and ADHD. The Diagnostic and Statistical Manual of Mental Illnesses, fifth edition (DSM-5) classifies dyslexia as the inability to link word patterns and pronunciations, affecting reading ability both internally and externally (5). People with dyscalculia struggle with ‘numeric and arithmetic’ concepts (6), whilst those with dysgraphia have lower abilities in fine motor coordination, than neurotypical individuals (7). Finally, ADHD is diagnosed when an individual exhibits reduced attentiveness and hyperactivity with actions of impulsivity (8).

Neurodevelopmental disorders are costly to both individuals and society. Disability adjusted life years (DALYs) are a quantification of years of full health lost to an individual. The DALYs associated with ADHD are 13.78 (9), and data suggest that the quality of life for individuals suffering from dyslexia, dyscalculia and dysgraphia was significantly lower in several domains than neurotypical subjects (10–12). Individuals suffering from a learning disorder are twofold less likely to gain a degree qualification after a full education. Such individuals are also more likely to be unemployed than neurotypical peers. (13, 14). In addition to this, poor mental health, emotional and behavioral issues are associated with specific learning disorders (15). The overall cost to an individual due to the burden of learning disorders can be categorized into medical (for diagnosis, treatment, and rehabilitation), non-medical (transport to appointments) and indirect costs (productivity losses, loss of earnings and academic enhancement measures). In 2012, the average annual cost to an individual with ADHD was estimated to be between £8,422.68 and £12,371.77, however, as more individuals are being diagnosed with multiple neurodevelopmental disorders, this burden is increasing (16). The direct, indirect, and intangible costs due to SLD in India were INR 5.94M, 29.26M, and 42.30M, respectively, with indirect costs making up 83.1%. Recent estimates show annual costs per child of £97,823.09 in India and total national expenditure of £569.48M in Australia (17) (18).

The current state of the art regarding the assessment of such disorders is varied. The National Institute for Health and Care Excellence in England (UK NICE) guidelines currently state that the diagnosis of ADHD is through assessing qualitatively the actions of an individual. There are dedicated methods such as the Strengths and Difficulties questionnaire or Conners’ rating scale (19). Conners’ rating scale is for the teacher, parent, and child self-report of conduct, self-care, and academic performance, and literacy level. SLD can be diagnosed by assessing intelligence, memory, and other cognitive skills of a child for basic reading, writing, and counting abilities using this rating scale (20). These assessments are currently carried out by an educational psychologist or a neurodevelopmental healthcare professional (21). This reduces the accessibility for individuals in low-income countries, remote areas, or those without access to adequate healthcare services being able to achieve a diagnosis and receive treatment. Early screening is indeed paramount, as it allows treatment to begin earlier, allowing the individual to receive support from an early age and reducing the costs to the individual and society (22). The current most effective treatment for ADHD is stimulants (methylphenidate and amphetamine) alongside behavioral therapy, boosting both dopamine and noradrenaline levels (23). Currently, there are no medications that treat dyslexia, dysgraphia, or dyscalculia. Instead, treatment includes educational accommodations, fine motor skill training and reading and writing lessons with trained professionals, both online and in person (24).

From the technological advances point of view, in recent years, it has been proposed that ‘serious gaming’ and ‘visual training’ can be used for the diagnosis and treatment of several learning disorders (e.g., dyslexia) (25) (26). Similarly, eye tracking has been identified as a potential diagnostics and rehabilitation tool in both ADHD (27, 28) and dyslexia (29, 30). In fact, eye-tracking technology delivers exceptional assessment abilities for neurodevelopmental disorders when its measurements exist independently from other measurement procedures. The objective and quantifiable capability of eye-tracking technology allows users to measure direct visual attention through oculomotor control to gauge gaze patterns, which commonly surface in ADHD and autism spectrum disorder as well as dyslexia patients. Furthermore, this technology allows clinicians to view subtle signs of abnormal thinking and vision processes which standard ways of testing might overlook (31, 32). Thus, early diagnostic symptoms may become evident. Moreover, eye-tracking monitoring now exists as a component within low-cost non-invasive screening arrangements, which allow its utilization in schools for early diagnosis and remote care. It also helps in rehabilitation by regularly watching how treatment is progressing. Additionally, these diagnostic capabilities of eye-tracking offer high accessibility along with scalability because the system provides real-time data while avoiding any need for verbal communication (33, 34). The analysis of eye movements has proven its standalone diagnostic capacity through reliable research findings indicating predictive and classification benefits that appear when using machine learning approaches or standalone measures (35). Research analysis of eye-tracking requires its own assessment because of its worthiness for presentation in this review (36).

Hence, this systematic literature review examines the available literature on the use of serious gaming and eye-tracking technologies for the early screening, diagnosis, monitoring, and treatment support of neurodevelopmental abnormalities in children. The research results will help guide the creation of a clinical decision support system that leverages AI and consumer-grade electronics (e.g., smartphones and tablets) to assist Paperbox Health’s endeavors. Paperbox Health is a start-up based in Italy that specialises in the detection and treatment of neurodevelopmental disorders using serious gaming.

## 2. Methods

The methodology followed the PRISMA statement for systematic literature reviews, ensuring a structured and transparent approach to reporting (see Figure 1) (37).

**Figure 1:**
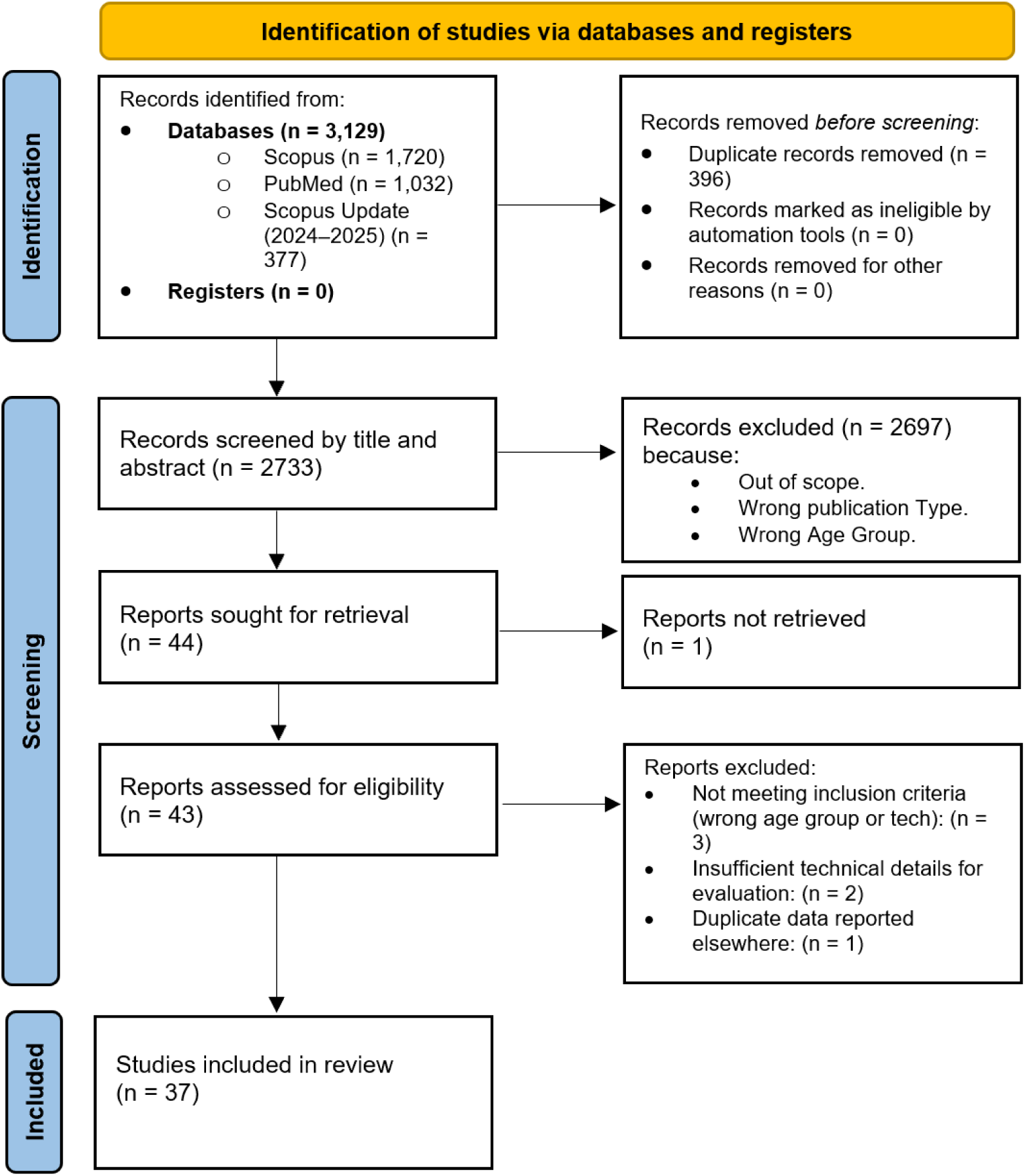
PRISMA flow diagram illustrating the screening and selection process for included studies.

The completed PRISMA checklist has been provided as Supplementary Material 1 to confirm adherence to established guidelines and facilitate reproducibility (37).

## 3. Search strategy

To perform this systematic review, a detailed and reproducible search strategy was developed. Meaningful keywords were identified through expert input and review of relevant literature. PubMed MeSH terms were used to structure part of the query and identify relevant synonyms. A focus group consisting of experts in biomedical engineering, medicine and speech and language therapy (SALT) finalized the search string. Boolean logical conjunctions (AND) along with disjunctions (OR) functioned to link search terms among five established categories such as neurodevelopmental diseases, digital gaming platforms, computational learning systems, child participants and assessment systems. All search strings employed for PubMed and Scopus investigation can be found in Supplementary Material 2. These search strings were inputted into Scopus and PUBMED, identifying research papers from January 2013 to February 2025.

## 4. Inclusion/exclusion criteria

After assessing existing reviews, we decided to continue with our systematic review effort, as we did not find any similar effort in the recent literature. We included only journal articles published in English from January 2013 to February 2025, related to screening neurodevelopmental disorders in children, possibly the studies that included children up to the age of 18 were included. Also studies with children already diagnosed with learning disorders were included, undergoing a technology-based intervention for the screening of neurodevelopmental disorders. We excluded reviews, conference papers, book chapters, editorials, notes, and those focusing only on epidemiology or using electroencephalography (EEG) or functional magnetic resonance imaging (fMRI) (see Table 1). Two authors independently appraised articles by title, abstract, and full text, with a third-party resolving discrepancy, if needed.

**Table 1.** Inclusion and exclusion criteria used in the systematic literature review.

| Inclusion criteria | Exclusion criteria |
| --- | --- |
| Publications from January 2013 to February 2025 |  |
| Original articles only | No conference papers, book chapters, reviews, editorials |
| Studies that included children up to the age of 18 were included. Also studies with children already diagnosed with learning disorders were included. | Studies did not include any children up to the age of 18. |
| English language | Any other language |
| Any study or manuscript describing how serious games, eye-tracking, machine learning or smartphone/tablet applications are applied for the screening, diagnosis, monitoring or treatment of neurodevelopmental disorders. | No use of electroencephalography (EEG) or functional magnetic resonance imaging (fMRI), no robots. |

## 5. Data extraction and quality appraisal

Relevant data was extracted and collated into an ad-hoc Excel spreadsheet. The quality appraisal was conducted using the MMAT tool (38), as the finalized selection included multiple study types. The MMAT tool allows for analysis of quantitative and qualitative studies.

## 6. Data synthesis

The narrative synthesis method (39) was used for data synthesis of the extracted data. For each paper, the type of disorder, technology and overall aims and conclusions of the study were identified and organised by the type of neurodevelopmental disorder. This information was used to generate discussions and future directions.

## 7. Results

### 7.1. Search outcome

This systematic literature review integrated PubMed and Scopus databases for data retrieval. The combined search returned 3,129 records (see Figure 1). After duplicate removal (396), the total number of screened records reached 2,733. After title and abstract screening, only 43 records seemed to be in line with the inclusion and exclusion criteria. Finally, a thorough full-text evaluation led us to the final set of 36 studies to be included in this review.

### 7.2. Data visualization and extraction

Table 2 contains the summary of the data extracted from the included studies. The following sections will report the findings by type of disorder and technology used.

**Table 2.** Summary of data extracted from the included studies, detailing the types of neurodevelopmental disorders investigated, the technologies employed in the research, the aims of each study, the number of participants and their age ranges, and the reported outcomes.

| Article No Citation. | Type of Disorder | Technology | Aim of the Study | Participants | Conclusion of the Study |
| --- | --- | --- | --- | --- | --- |
| Apiquian, 2020 (40) | ADHD | Video game | To determine whether their video game (Chefmania) was a useful tool in | 266 participants, aged 6 - 12 | The intervention, consisting of Chefmania scores, showed that children with |
|  |  |  | screening ADHD |  | ADHD, aged 6 - 12 performed poorly, suggesting the potential of Chefmania as a valid tool for assessing cognition in schools. |
| Fernández-Martín et al., 2024 (32) | ADHD | Educational video games | To explore modern techniques which can be used to create data models for ADHD based on ecological assessments of attention problems and distractibility combined with hyperactivity indicators, using virtual reality. | 110 participants, with a mean age of 9.47 (standard deviation of 2.93) | The research produced distinctive behavioral profiles for improving the delivery of ADHD interventions. |
| García-Baos et al., 2019 (41) | ADHD | Eye-tracking video game | To assess the therapeutic potential of an eye-tracking game (RECOGNeyes) for training visual attention and oculomotor control in children with ADHD | 28 participants, aged 8–15, previously diagnosed with ADHD (DSM-5 criteria) | The eye-tracking group showed significant improvements in reaction time, gaze fixation, and impulsivity control, suggesting RECOGNeyes may be a useful non-pharmacologic tool for ADHD training. |
| García-Redondo et al., 2019 (42) | ADHD | Educational video games | To analyze the effects of a serious game based on multiple intelligences on attention in students with ADHD and SLD, using performance and observation measures. | 44 students, aged 6 - 16 | The intervention, consisting of 28 sessions using educational video games, resulted in a significant improvement in attention performance, particularly visual attention, suggesting the potential of serious games in addressing attention issues in students with learning disabilities. |
| Garcia-Zapirain et al., 2017 (43) | ADHD | Eye-tracker and hand gesture sensors | To evaluate a rehabilitation system using physiological sensors | 19 participants, aged 10 | The system, based on user feedback, showed high effectiveness in rehabilitation, suggesting its potential in improving rehabilitation outcomes. |
| Heller, 2013 (44) | ADHD | Machine Learning, video game | To test the accuracy of a game implementing machine learning at identifying ADHD. | 52 participants, aged 6 - 17 | The game was able to accurately detect combined type ADHD 75% of the time and inattentive type 78% of the time demonstrating the potential to create a game to detect patterns for ADHD and potentially other mental disorders. |
| Oh et al., 2024 (35) | ADHD | Eye-tracking | The research investigates the effectiveness of AttnKare-D, an artificial intelligence software that evaluates Attention Disorders using virtual reality evaluations. | 20 children aged 6–12 (15 ADHD, 5 control) | Virtual Reality (VR) and AI integration improved diagnostic accuracy, simulating real-life environments. |
| Slobodin et al., 2020 (45) | ADHD | Data Collection using MOXO-CPT | To use a machine-based learning model to predict ADHD in children | 458 participants, aged 6 - 12 | The MOXO-CPT assessment predicted ADHD with high accuracy, quicker and more cost effective than the current ADHD diagnosis showing 87% accuracy, 89% sensitivity and 84% specificity. |
| Takahashi et al., 2024 (31) | ADHD | 3D Video Game, and Eye-tracking | To investigate if ADHD children | 33 participants | The game metrics showed parallel |
|  |  |  | can be assessed through executive function performance on a 3D action puzzle video game. | aged 8–21 | results to neuropsychological test results which suggests that the game can evaluate executive functioning in patients with ADHD. |
| Toki et al., 2024 (36) | ADHD, Neurological Disorders | Educational video games | To explore machine learning methods to forecast possible neurodevelopmental disorders among children. | 473 participants for the model; 184 participants with an average age of 7 for the predictive analysis. | ML models demonstrated good prediction capabilities for early diagnosis, based on cognitive features. |
| Türkan et al., 2016 (46) | ADHD | Video-based eye-tracking | To investigate oculomotor abnormalities in ADHD | 48 participants (24 children with ADHD and 24 typically developing children, mean age = 8 years, 10 months). | The intervention showed that children with ADHD, aged 6 - 17, had shorter fixation periods compared to neurotypical peers, suggesting differences in attention performance. |
| Yoo et al., 2024 (33) | ADHD | Eye-tracking | To explore the creation of an eye-tracking feature-based machine learning screening system for ADHD. | 135 participants (56 with ADHD and 79 typically developing children), with an average age of 8.6 years old. | The research team achieved 76.3% accuracy using eye-tracking-only features, which demonstrates the potential of ML models for ADHD screening. |
| Sweere et al., 2022 (47) | ADHD, Neurological Disorders | Eye-tracking and manual response | To investigate distraction effects during reaction tasks | 177 participants, aged 6 - 17 | The intervention, based on eye movement data, demonstrated increased distractibility in ADHD/neurological groups, with no significant latency differences, suggesting potential insights into attention performance in these groups. |
| Perochon, 2023 (48) | Autism and ADHD | Tablet based game | To use extracted touch features from a bubble popping game to differentiate between neurotypical and neurodivergent children | 233 participants, aged 1.5 - 10 | The intervention, involving touch-based games, showed that autistic children with co-existing ADHD took longer to pop the bubble and had a longer touch length on average, suggesting the potential of touch-based games as an efficient approach to screen autism and ADHD. |
| Van Viersen et al., 2013 (49) | Dyscalculia | Eye-tracking | To examine abnormal estimation patterns in dyscalculia | 11 participants (a 9-year-old girl with developmental dyscalculia and 10 typically developing children). | The intervention, using eye-tracking, effectively identified abnormal estimation patterns in dyscalculia, suggesting its potential as a tool for diagnosing dyscalculia. |
| Piazzalunga et al., 2023 (50) | Dysgraphia | Visual perception games, and and eye-tracking device | To investigate the role of visual perception in handwriting skills | 53 participants, aged approx. 8 | The intervention showed that children with dysgraphia displayed more inattentiveness and indecisiveness, suggesting the potential of this tool to predict risks in handwriting with good accuracy. |
| Alt et al, 2017 (51) | Dyslexia | Computer-based word learning games | To use word learning games for identifying strengths and weaknesses in children with | 184 participants, aged 7 - 8 | The study found word learning deficits in children with dyslexia across all manipulations and |
|  |  |  | dyslexia by measuring their ability to link names and objects. Analyses of covariance and nonverbal intelligence scores assessed phonological-visual linking, mispronunciation detection, naming, visual difference decision, and visual feature recall. |  | most task-manipulation combinations, especially in phonological tasks. Visuospatial manipulations showed mixed effects. |
| Gomolka et al., 2024 [74] | Dyslexia | Eye-tracking | To diagnose dyslexia in early school-aged children using eye-tracking and LSTM networks. | 145 children aged 7-10 | The Model displayed 97.7% accuracy making it suitable for diagnosing dyslexia in its early stages. |
| Ikermane, 2023 (52) | Dyslexia | Webcam-based eye-tracking | To study reading behavior using Arabic text configurations | 61 participants, aged 9 - 13 | The intervention, using webcam-based eye-tracking, showed that dyslexic individuals exhibited longer fixations and reading periods, suggesting its diagnostic potential for dyslexia. |
| Jafarlou et al., 2017 (30) | Dyslexia | Videonystagmography (VNG) | To assess oculomotor rehabilitation in dyslexia | 50 participants aged 8-12 years (30 dyslexic children and 20 typical readers). | The intervention, involving oculomotor rehabilitation, improved eye movement patterns, suggesting its potential to aid reading and visual search abilities. |
| Jafarlou, et al, 2021 (53) | Dyslexia | Eye-movement tracking | To evaluate the effectiveness of oculomotor rehabilitation on the cognitive performance of | 50 participants aged 8–12 years (30 dyslexic children and | The intervention showed a significant difference between children with and without |
|  |  |  | dyslexic children. | 20 non-dyslexic children). | dyslexia in signs of oculomotor impairment, suggesting that early diagnosis of eye movement disabilities can improve cognitive performance in affected children. |
| Korneev et al., 2018 (54) | Dyslexia | EyeLink 1000 eye-tracking device | To investigate reading mechanisms in young readers | 36 participants, aged approx. 9 | The intervention showed that longer words caused more fixations, suggesting that proficient readers used lexical strategies more effectively. |
| Larco et al., 2021 (55) | Dyslexia | Web and app-based learning game | To create web and mobile apps to provide learning resources to dyslexic children aged 7-10 years. | 25 participants, aged 7 - 10 | The intervention, involving the 'Helpdy' app, showed improvement in the symptoms of dyslexia in children, suggesting its potential as a tool for supporting dyslexia management. |
| Masulli et al., 2018 (56) | Dyslexia | Video-based eye-tracking | To investigate effects of text format on eye movements | 45 participants aged 7-12 years (15 dyslexic, 15 reading-age-matched non-dyslexic, and 15 chronological-age-matched non-dyslexic). | The intervention showed that dyslexic children had longer fixation durations, with font size and spacing reducing fixation durations, suggesting their potential in improving reading performance for dyslexic individuals. |
| Moiroud et al., 2018 (57) | Dyslexia | Eye-tracking with DEM Test | To analyze oculomotor behavior in dyslexic children | 39 participants aged 7-12 years (13 dyslexic, 13 chronological-age- | The intervention showed that dyslexic children had longer fixation durations, with font size and spacing reducing |
|  |  |  |  | matched non-dyslexic, and 13 reading-age-matched non-dyslexic). | fixation durations, suggesting their potential in improving reading performance for dyslexic individuals. |
| Nilsson Benfatto et al., 2016 (58) | Dyslexia | Video-based eye-tracking | To predict dyslexia risk using eye-tracking data | 185 participants aged 9–10 years (97 high-risk with early word decoding difficulties and 88 low-risk control). | The intervention, using eye-tracking, predicted dyslexia risk with 96% accuracy, suggesting its potential as a reliable tool for early dyslexia detection. |
| Prabha & Bhargavi, 2020 (59) | Dyslexia | Machine learning with eye-tracking | To compare dyslexic vs. non-dyslexic traits using ML models | 187 children aged 9-10 years old, of which 97 were dyslexic and 88 non-dyslexics | The PSO-SVM Hybrid Kernel achieved 95.6% accuracy in identifying dyslexia traits, suggesting its potential as an effective tool for dyslexia detection. |
| Prabha & Bhargavi, 2022 (60) | Dyslexia | Machine learning with eye-tracking | To develop screening tool for dyslexia diagnostics | 187 children aged 9-10 years old, of which 97 were dyslexic and 88 non-dyslexics | The intervention validated the PSO-SVM Hybrid Kernel as an effective screening tool for schools, suggesting its potential in identifying dyslexia traits. |
| Rauschenberger, 2022 (61) | Dyslexia | Web-game | To develop a game for universal screening of dyslexia | 313 participants aged 5-10 years (116 with dyslexia, 197 non-dyslexic). | The intervention showed seven separate variables with significant differences between those with and without dyslexia, suggesting that this approach can optimize resources for detecting dyslexia, though it needs to expand training data. |
| Rello et al, 2020 (62) | Dyslexia | Online gamified test | To design an online gamified test and a predictive machine learning model for dyslexia detection in Spanish speakers. | 3,644 participants (general age group unspecified) and 1,300 participants aged 12 years or older in a follow-up study. | The study correctly detected 80% of participants with dyslexia among over 3,600 participants, suggesting the potential of the online screening tool, used by over 200,000 people, for early detection and prevention of dyslexia-related challenges. |
| Vajs and Papi, 2023 (29) | Dyslexia | Remote eye-tracking (SMI RED-m) | To analyze eye movements while reading with color configurations | 30 participants aged school-age (15 dyslexic and 15 neurotypical ). | The intervention, using eye-tracking data, effectively differentiated dyslexic and control groups with high accuracy, suggesting its potential as a reliable tool for dyslexia detection. |
| Vajs et al., 2022 (63) | Dyslexia | Spatiotemporal eye-tracking | To differentiate dyslexic and non-dyslexic readers | 30 participants aged 7–13 years (15 dyslexic and 15 non-dyslexic). | The intervention, using spatiotemporal features, achieved 94% accuracy in distinguishing dyslexic readers, suggesting its potential as an effective tool for dyslexia identification. |
| Van de Ven et al, 2017 (64) | Dyslexia | Reading Game (Letter Prince) | To measure the impact of a multicomponent reading game on the development of reading skills and motivation in 60 first grade Dutch schoolchildren with special | 60 participants aged primary school age (Dutch children with special educational needs). | The short intervention (9 × 15 min) improved pseudoword and text reading fluency, with early intervention reducing reading time and late intervention increasing |
|  |  |  | educational needs. |  | pseudoword identification, showing no impact on reading motivation. |
| Bonifacci et al., 2023 (65) | Dyslexia, Preterm Birth | Video-based eye-tracking | To compare eye movements in dyslexia and preterm birth | 78 participants, aged 10.5 years (preterm, dyslexic, and typically developing children). | The intervention showed that dyslexic children had increased fixations and smaller saccades, while preterm children exhibited reduced reading speed, suggesting potential differences in reading performance. |
| Schmitt, 2018 (66) | N/A | Web-based educational video games | To evaluate the effectiveness of an educational website in promoting the literacy skills of young children via a randomised trial over 8 weeks (about 2 months). | 136 participants, aged 4 - 7 | The intervention, using the website, showed that children had better literacy results after assessment compared to the control group, suggesting its potential to improve literacy outcomes. |
| Peters et al., 2020 (67) | Reading Difficulties | Eye-tracking | To examine the relationship between RAN and reading skills | 93 participants, aged 5 - 8 | The intervention, using fixation metrics, predicted RAN performance and differentiated between reading difficulties and neurotypical peers, suggesting its potential for identifying reading challenges. |
| Niemela, 2020 (68) | Reading disability | Serious Games and Clustering | To implement a new clustering-based approach for identifying different profiles of serious | 136 participants, with varying age (specific age not mentioned), all learners of GraphoLear | The intervention showed that it is possible to identify different types of learners, suggesting its potential for tailoring educational |
|  |  |  |  | n, analyzed using a clustering-based profiling method to detect reading difficulties. | approaches. |

### 7.3. Dyslexia

Dyslexia was the most extensively studied disorder, featuring in 18/37 (48.6%) of the included studies, with technologies such as eye-tracking, educational games, and machine-learning-based software receiving significant attention.

Eye-tracking was a particularly prominent tool, offering insights into the reading behaviors and oculomotor patterns of individuals with dyslexia. For instance, Vajs et al. (29, 63) demonstrated that spatiotemporal features (e.g., fixation intersection coefficient and fixation fractal dimension) extracted through eye-tracking could distinguish dyslexic from non-dyslexic readers with 94% accuracy, based on a group of 30 children. Similarly, Nilsson Benfatto et al (29, 58, 63). achieved 96% accuracy in predicting dyslexia risk using video-based eye-tracking, based on 97 high-risk subjects and a control group of 88 low-risk subjects, while Korneev et al. (2018) (54) investigated the reading mechanisms of 36 second-grade pupils using eye tracking. Their results highlighted a close relation between the level of the reading skill and the oculomotor activity. Additionally, Bonifacci et al. (2023) (65) utilized eye-tracking to compare reading behaviors among children with dyslexia, those with typical development, and those born preterm, identifying distinct oculomotor patterns in both groups, such as more fixations and more frequent and smaller saccades for the groups affected by dyslexia. Prabha and Bhargavi et al. (2020) (59) focused on eye-tracking as a means of analysing cognitive processing in dyslexics using machine learning models to compare dyslexic versus non-dyslexic traits. Specifically, fixations and saccades were detected and evaluated with machine learning algorithms. Gomolka et al. (2024) [74] implemented a new method for early dyslexia testing of school children through eye-tracking analysis by an LSTM-based deep learning model. The objective of the research focused on identifying dyslexic versus non-dyslexic children through oculomotor analysis of their reading tasks. Their research exhibited outstanding diagnostics outcomes, which confirmed the power of artificial intelligence models combined with eye-tracking technology for early dyslexia detection at educational institutions. Particle Swarm Optimization based SVM Hybrid Kernel was the best performing model, reaching 95.6% of accuracy. The same authors, in 2022, Prabha and Bhargavi (2022) (60) conducted a subsequent study further validating the Particle Swarm Optimisation based SVM Hybrid Kernel model on 187 kids, resulting in a proposed screening tool for the diagnosis of dyslexia in schools. Also, Masulli et al. (2018) (56) studied eye movements, but aimed to compare the changes in eye movement between 15 children with and 15 children without dyslexia by reading four lines of text each with differing font sizes and word spacing. They found that increased font size and spacing caused a reduction in the duration of eye fixation and increased prosaccade amplitude in all children. They also found that the duration of fixation was significantly longer in dyslexic children. Jafarlou et al. (2017, 2021) (30, 53) evaluated the differences between eye movement patterns in Iranian dyslexic children compared to non-dyslexic children, with the latter presenting lower gain in pursuit and optokinetic tests, and increased latency with decreased accuracy in the saccade test. In their second study of 2021, they also assessed the effectiveness and impact of oculomotor rehabilitation on children with developmental dyslexia (53). Moiroud et al. (2018) (57) explored eye movement recordings during the Developmental Eye Movement test in dyslexic and non-dyslexic children. They found that children with dyslexia and non-dyslexic children of equivalent reading age have significant longer fixation time and take longer to read than non-dyslexic children of similar chronological age. A significant correlation was also found between the fixation time and the number of words read in one minute with the total reading time. Similarly, Ikermane and El Mouatasim (2023) (52) analysed eye-tracking data of 61 subjects who were given Arabic normal text materials with various text configurations such as font-sizes, character spacing, and font family to analyse their reading behaviour. They found also that the eye motions of those with dyslexia differ from those of neurotypical readers. People with dyslexia tend to have longer and more frequent fixations, and longer reading time. Results confirmed that webcam-based eye-tracking techniques can be used as an assessing tool for investigating the student’s reading performance and the potential risk of dyslexia. Finally, other innovative approaches included the use of different color configurations combined with eye-tracking. Vajs and Papi (2023) (29) analysed the eye-tracking data from 15 dyslexic and 15 neurotypical Serbian school-age children who read 13 text segments presented on different colour configurations (29). They trained and tested a machine learning algorithm obtaining accuracies greater than 87%, and showing that the color configuration, despite it may influence each subject differently, does not affect the outcome.

Educational games were also widely explored as tools for both diagnosis and intervention. Rauschenberger et al. (2022) (61) developed a web-based game (MusVis) which was able to observe significant differences between children with and without dyslexia, relying on seven significant variables including duration round, duration interaction and average click time. Larco et al. (2021) presented their app (Helpdy) which offers engaging educational games like *El Ahorcado* (Hangman) and *Memoria* (Memory) to enhance cognitive skills such as memory, concentration, and word recognition (55). The app also features two interactive pre-diagnosis games: one for phonological dyslexia (vowel placement to form words) and another for visual dyslexia (word selection tasks) (55). Rello et al. (2020) (62) combined gamified online tests with machine learning models to predict dyslexia risk, achieving 80% success in Spanish-speaking populations, based on a study of more than 3,600 participants. Their game was designed based on typical mistakes made by Spanish people with dyslexia and on dyslexia-related indicators such as Language Skills, Working Memory, Executive Functions and Perceptual Processes (62).

Furthermore, van de Ven et al. (2017) (64) proposed an adaptive learning platform driven by reinforcement learning to optimize reading practice for children with dyslexia, showing significant improvement in reading fluency (64). Specifically, they found a significant improvement in pseudoword naming and reading time after the use of a computer game. Alt et al. (2017) (51) used word learning games and demonstrated that children with dyslexia face significant word learning deficits, especially when working memory demands are high, highlighting the interplay between cognitive load and language acquisition in dyslexic populations. However, the software they used was limited to a dichotomic choice (Yes VS No), with a 50% chance for the kid to guess the correct answer (51).

### 7.4. ADHD

ADHD was another well-represented disorder, featuring in 14/37 (37.8%) of the included studies, which explored video games, machine learning, eye-tracking, and physiological sensors as diagnostic and intervention tools. Several studies employed video games to assess cognitive abilities and attention performance. For example, Apiquian et al. (2020) (40) used the “Chefmania” game assessing attention, memory, and impairment in executive functions to compare children with and without ADHD, showing a statistically significant difference, highlighting that this may be a plausible screening technique in this remit. They also noted that artificial intelligence could be used to allow adaptations for subjects with cognitive impairments (40). Similarly, Garcia-Redondo et al. (2019) [45] demonstrated the potential of educational video games based on the Tree of Intelligences method (e.g., “Boogies Academy”) to improve attention in students with ADHD and specific learning disabilities, showing significant gains in visual attention after 28 sessions (42). This shows that computer intervention could not only act as a promising future diagnostic tool for ADHD but also offer a therapeutic advantage in the treatment of ADHD. Perochon (2023) (48) introduced a tablet-based bubble popping game aimed at assessing visual-motor skills in children with ADHD and comorbid conditions such as autism. The study revealed distinct visual-motor patterns, including longer response times and higher error rates, indicating the potential of touch-sensitive games for early detection of ADHD-related symptoms in children also affected by autism (48). Modern advancements in digital technology support several new studies that use them to improve ADHD detection. The assessment of executive functions in ADHD patients through 3D-video games was investigated by Takahashi et al. (2024) [71]. This research with 33 participants aged 8–21 demonstrated that in-game and executive functioning test results had a strong relationship which supports digital tools that mimic real-world environments for assessing ADHD.

Machine learning approaches were also applied to ADHD diagnostics. Heller et al. (2013) (44) tested a game (“Groundskeeper”) integrating machine learning algorithms and demonstrated high accuracy in identifying ADHD subtypes, detecting combined-type ADHD with 75% accuracy and inattentive type with 78% accuracy. Similarly, Slobodin et al. (2020) (45) employed a machine-based learning model within the MOXO-CPT framework inclusive of visual and auditory stimuli distractors, achieving 87% accuracy, 89% sensitivity, and 84% specificity in ADHD prediction.

Eye-tracking technologies provided additional insights into ADHD-related behaviors. Türkan et al. (2016) (46) investigated oculomotor abnormalities in children with ADHD, identifying shorter fixation periods and higher distractibility compared to neurotypical peers. Sweere et al. (2022) (47) further demonstrated how eye movement data from a visual reaction time task (“iDistrack”) can be used to quantify distractibility in ADHD populations, highlighting their heightened susceptibility to external stimuli. However, they also noted that the expected differences in manual press latencies between the ADHD and non-ADHD group were not observed (47). Garcia-Baos et al. (2019) carried out a large-scale randomised control trial of interactive eye track game (RECOGNeyes) that aimed at training ocular motor control and visual attention in children with ADHD. During the study 28 participants were tested (ages 8-15) and the results proved that the participants who played the eye-controlled game performed better in terms of reaction time, fixation length of their gaze as well as being less impulsive as compared to the control groups who used mouse tracking. The results highlight the therapeutic effects of gaze-contingent video games in the improvement of visual attention systems and the oculomotor controls in ADHD groups of people (41). A portable eye-tracking system merged with machine learning algorithms became available for ADHD screening as described by Yoo et al. (2024) [73]. The research examined eye-tracking biomarkers among 56 participants aged 6–12 which showed movement latency and fixation time allowed researchers to achieve 76.3% accuracy in screening results. Studied results confirmed that cost-effective diagnostic systems employing eye-tracking technologies can work properly in medical environments. The research work of Oh et al. (2024) [75] analyzed classroom simulations implemented in virtual reality with artificial intelligence elements during which 40 children aged 7–13 took part. The analysis through deep learning processing of their eye-tracking and behavioral activities showed early indications for real-world application success in ADHD prediction.

Other innovative tools included multimodal systems that combine various sensors and interfaces. For instance, Garcia-Zapirain et al. (2017) (43) incorporated hand gesture recognition sensors and an eye tracker into a rehabilitation platform designed for ADHD intervention, receiving positive feedback on its usability and effectiveness in improving attention and motor coordination. A machine learning system by Toki et al. (2024) [76] analyzed behavioral along with environmental data from 78 children between ages 6 and 12 to create the model. The research confirmed that AI models can make precise predictions about neurodevelopmental risks due to their general applicability. A virtual reality-based continuous performance test (AULA) helps Fernández-Martín et al. (2024) [72] evaluated attention capabilities as well as hyperactivity and distractibility in 110 children between the ages of 6–16. New ADHD profiles emerged through cluster analysis which extended beyond DSM-5 subtypes because head motion and response time performances revealed themselves as effective clinical diagnosis tools.

### 7.5. Dyscalculia and Dysgraphia

Dyscalculia and dysgraphia, although less frequently studied (2/37, 5.4%), have also shown potential for innovative technological interventions. While current data remains preliminary, there is compelling evidence to support the efficacy of these approaches. Research on dyscalculia has highlighted difficulties in spatial cognition and numerical representation, effectively identified through real-time eye-tracking analyses. For example, Van Viersen et al. (2013) (49) employed eye-tracking to examine estimation patterns in individuals with dyscalculia, identifying distinct abnormalities in the processing of mathematical tasks. These findings underscore the utility of eye-tracking for detecting specific markers associated with dyscalculia, differentiating them from control groups (49). Similarly, technology has demonstrated promise in the early detection and intervention of dysgraphia. Dysgraphia is often diagnosed belatedly since its identification typically occurs only after handwriting skills are fully developed (49). Technological tools could therefore support schools in the early screening of this SLD by analyzing indicators such as visual perception skills and eye movements (49). These tools provide opportunities to strengthen impaired abilities at an earlier stage (49). Piazzalunga et al. (2023) (50) investigated the role of visual perception games and eye-tracking in children with dysgraphia, finding increased inattentiveness and indecisiveness during handwriting tasks. Their study demonstrated that these tools can reasonably predict handwriting difficulties and associated risks, even before children fully develop writing skills (50). In conclusion, these tools show strong potential for early screening of dysgraphia and offer a cost-effective solution with significant benefits (50). Additionally, they could serve as a method to explore the role of specific abilities in the development of handwriting (50). Despite their promise, the current levels of accuracy and precision remain insufficient (50). Longitudinal studies are essential to improve the robustness and generalizability of the findings, paving the way for more reliable and impactful interventions that address fine motor and visual-spatial processing deficits (50, 69).

### 7.6. General Reading Difficulties

General studies on reading difficulties (2/37, 5.4%) highlight that patterns and trends can be identified in players’ results when engaging with educational video games. Research employing serious games, eye-tracking, and clustering techniques has demonstrated their potential in analyzing and enhancing reading performance, with educational profiles emerging from broader assessments (69). Niemelä (2020) (68) applied clustering algorithms to GraphoLearn game log data, identifying six distinct learner profiles among over 1,600 children. These profiles, characterized by specific errors such as confusion between phonetically or visually similar letters, showcase the potential of serious games like GraphoLearn in supporting reading acquisition within transparent writing systems (68). Learners with higher initial error rates displayed notable progress, proving the game’s effectiveness for severe reading difficulties (68). While the method enables early identification of children at risk for reading disabilities and supports targeted, personalized interventions, its reliance on early-stage data limits insights into long-term skill development (68). Future research should focus on expanding datasets, analyzing variables such as syllables and words, and refining clustering algorithms to deepen understanding and enhance game design (68). Additionally, Peters et al. (2020) (67) investigated the relationship between rapid automatized naming (RAN) and reading skills, demonstrating that fixation metrics from eye-tracking can predict RAN performance and distinguish between children with and without reading difficulties. This highlights the potential of eye-tracking in identifying early indicators of reading challenges, facilitating earlier and more effective interventions (67).

These findings underscore the importance of integrating serious games and technology such as eye tracking to personalize educational interventions and identify those at possibly affected by learning disorders early.

### 7.7. Technologies and Techniques

Across all the studies that were examined, the most common technology applied was eye-tracking in 21 (56.75%), followed by educational/serious games in 14 studies (37.38%). Machine learning (ML) techniques were identified in 7 studies (18.9%), most often in combination with other equipment like eye-tracking or gamified systems. The focus of this type of research was primarily on ADHD (14 studies) and dyslexia (18 studies), and to a lesser extent on dyscalculia, dysgraphia, or other neurological disorders. Furthermore Eye-tracking was the most generic technology used, most importantly for early identification, as it could track fixations without subject control. Additionally, game-based technologies were used extensively for screening purposes and cognitive training, generally boosting motivation and motivation in children. Moreover, ML-based technologies stepped in to recognize predictive models with high precision in diagnosing neurodevelopmental disorders. Overall, Table 2 reflects a transdisciplinary tendency for integrating interactive environments (e.g., VR, educational games) with computational smartness (e.g., ML, biometric sensing) to deal with the multidimensionality of children’s learning and attention disorders.

### 7.8. Quality Appraisal

The MMAT quality analysis results are shown in the table in Supplementary Material 3. A total of 36 studies were included in the review, all of which had identifiable and specific research questions aligned with the available data. Among these, two studies had unclear details regarding the population, and none provided adequate control for confounders due to missing or inadequate follow-up outcomes. This limitation may affect the correct interpretation of the data and introduce potential bias. Additionally, one randomized controlled trial was included, but it remains unclear whether the two groups were comparable at the beginning of the study, whether blinding was applied to the assessor, and whether participants adhered to the intervention.

## 8. Discussions

### 8.1. Principal Findings and Comparison to Prior Work

This review underscores the significant advancements in leveraging innovative technologies to address SLDs such as dyslexia, ADHD, dyscalculia, dysgraphia, and general reading difficulties (70, 71). Across these domains, tools like eye-tracking, educational games, and machine learning have emerged as powerful instruments for enhancing diagnosis, understanding, and intervention (8, 23, 27, 70, 71).

Dyslexia was the most extensively studied disorder, with eye-tracking proving particularly transformative (24, 72). This technology provided critical insights into reading behaviors and oculomotor patterns, offering high diagnostic accuracy and deepening understanding of the reading challenges faced by individuals with dyslexia (73). The integration of machine learning with eye-tracking has shown promise for scalable diagnostic tools, while text configuration adjustments and color-based analyses have highlighted opportunities for tailored interventions (10, 20, 25, 73). Educational games also emerged as engaging tools for both diagnosis and therapy, demonstrating their capacity to address cognitive deficits and enhance reading fluency through gamified experiences (72).

A similar range of technologies was used for ADHD (45). Video games offered an engaging medium for assessing cognitive abilities and attention, while also showing therapeutic benefits (27, 43, 66). Machine learning approaches demonstrated high accuracy in diagnosing ADHD subtypes, enhancing precision and paving the way for personalized interventions thanks to clustering techniques (44, 45, 74). Eye-tracking studies revealed characteristic oculomotor behaviors, providing valuable insights into distractibility and attentional deficits typical of individuals with ADHD (30). Combining eye-tracking with additional input sources, such as hand gestures, sensibly enhance the accuracy of the prediction suggesting for a multimodal approach (61). However, increasing the complexity of the tools used subsequently, decreased the accessibility.

Although less frequently studied, dyscalculia and dysgraphia have shown benefit from emerging technologies (6, 12, 49). Eye-tracking was effective in identifying spatial and numerical representation deficits in dyscalculia, while tools analyzing visual perception and handwriting behaviors showed promise for early dysgraphia screening (49). These findings highlight the potential of technology to address challenges in mathematical and writing skills at an earlier stage (6).

### 8.2. Strengths & Limitations

For general reading difficulties, serious games and eye-tracking demonstrated their value in identifying reading patterns and supporting personalized interventions, showing a great potential in tasks like clustering and prediction (5, 54, 67, 75, 76). Tools that combined gamified learning with advanced data analysis helped profile learners and provided targeted support, emphasizing the role of technology in early detection and tailored educational strategies (6, 13, 17, 21, 64, 77).

New scientific investigations strengthen the importance of eye-tracking in combination with AI-based innovations for ADHD assessment. Researchers demonstrated the ability of virtual reality in combination with portable eye trackers and 3D games to assess attention processes accurately according to multiple studies [71–76]. The combination of multimodal systems with machine learning algorithms has achieved strong diagnostic success and practical implementations in clinical as well as school environments subsequently emphasizing the significance of these technologies for personalized scalable intervention programs.

Furthermore, from the methodological point of view, this study has several limitations. Firstly, the search strategy was confined to Scopus and PUBMED, which might have excluded some evidence available in other databases (e.g., Web of Science). Nevertheless, Scopus and PUBMED are among the largest repositories currently available, and we deemed them adequate for our objectives. Secondly, only articles written in English were included, potentially omitting some relevant information. However, non-English articles constituted a negligible proportion of the total. Moreover, although one of our inclusion criteria specified studies involving children up to the age of 18, we occasionally deemed it relevant to include studies that extended participant recruitment to individuals up to the age of 21, even if they primarily focused on children up to 18 (e.g (31)). Finally, the potential for publication bias cannot be overlooked, as it affects all reviews and relies on published literature, which may not reflect all conducted research. Our quality assessment analysis, conducted using the MMAT tool, provides additional insights into the quality of the studies included.

## 9. Implications for Future Research

Eye-tracking and machine learning have demonstrated remarkable accuracy in controlled environments, but broader validation using larger datasets and more diverse populations is necessary (31–36). Future research should prioritize integrating these technologies into comprehensive frameworks that combine diagnostic precision with tailored interventions. This approach would empower educators and clinicians to more effectively support individuals with SLDs. In addition, it is noteworthy that there is a lack of robust longitudinal studies, which are essential to observe the progression of at-risk groups over time, from assessment to diagnosis. This would help highlight how early intervention improves children’s skills and quality of life over time and identify replicable patterns for early intervention strategies. The road to early screening and intervention for learning disorders is long, but the key findings of this review highlight possible solution paradigms, which can be used to streamline the existing processes and move towards an ever more digital approach, making the process quicker and easier to attain. After all, this is one of the core aims of Paperbox Health, which is looking after the design and validation of serious gaming software aimed at the early diagnosis of neurodevelopmental disorders, leveraging consumer-grade electronics (e.g., smartphones and tablets) and artificial intelligence.

## 10. Conclusions

The findings of this review underscore the significant potential of emerging technologies to transform the diagnosis, understanding, and treatment of SLDs. Nonetheless, critical challenges remain, including scalability, generalizability, and the long-term impact of these innovations, which warrant further exploration. Despite the presence in the scientific literature of this cutting-edge research, little to none has yet been translated into clinical practice. Speech therapists and specialists of neurodevelopmental disorders still recur to paper-based tests and examinations, which are not very efficient in terms of constant monitoring and vary significantly from country to country. With Health 4.0 under way, and Health 5.0 being the future step, digitization and personalization are key pillars for healthcare (78). This is well-aligned with the UK NHS Long Term Plan aims of improving access to services, enhancing patient experience, supporting clinical decision-making, and enabling more efficient and effective delivery of care (70, 79).

## 11. Abbreviations

ADHD: Attention Deficit Hyperactivity Disorder
DALYs: Disability Adjusted Life Years
DSM-5: Diagnostic and Statistical Manual of Mental Disorders, Fifth Edition
EEG: Electroencephalography
fMRI: Functional Magnetic Resonance Imaging
MMAT: Mixed Methods Appraisal Tool
NHS: National Health Service
NICE: National Institute for Health and Care Excellence
PRISMA: Preferred Reporting Items for Systematic Reviews and Meta-Analyses
PSO: Particle Swarm Optimization
RAN: Rapid Automatized Naming
SD: Standard Deviation
SLD: Specific Learning Disorder
SUS: system usability scale
QUIS: questionnaire for user interaction satisfaction
VNG: Videonystagmography

## Data Availability

All data produced in the present work are contained within the manuscript.

## 12. Declarations

### 12.1. Ethical Approval

Not applicable.

### 12.2. Informed Consent

Not applicable.

### 12.3. Statement Regarding Research Involving Human Participants and/or Animals

Not applicable.

### 12.4. Consent to Participate

Not applicable.

### 12.5. Consent to Publish

Not applicable.

### 12.6. Funding

This project was funded by a Warwick Industrial Fellowship agreement with Paperbox Health. C.H. was supported by the Beacon Academy, University of Warwick.

### 12.7. Author’s Contribution

Conceptualization: DP, GP, SS

Methodology: DP

Software: MFS, CH, CC, EP, RF

Validation: MFS, CH, CC, EP, RF

Verification: MFS, CH, CC, EP, RF

Formal analysis: MFS, CH, CC, EP, RF, SS

Investigation: MFS, CH, CC, EP, RF, SS, GP, DP

Resources: DP, GP

Data Curation: MFS, CH

Writing - Original Draft: MFS, CH, CC, EP, RF, SS, GP, DP

Writing - Review & Editing: MFS, CH, CC, EP, RF, SS, GP, DP

Visualization: MFS, CC

Supervision: GP, DP

Project administration: GP, DP

Funding acquisition: GP, DP

### 12.8. Competing Interests

Giacomo Pratesi, one of the authors, is the founder and Chief Technology Officer (CTO) of Paperbox Health, a company that specializes in the detection and treatment of neurodevelopmental disorders using serious gaming.

### 12.9. Availability of Data and Materials

All data generated or analyzed during this study are included in this published article (and its supplementary information files).

***Supplementary Material 1:*** *PRISMA Checklist*.

***Supplementary Material 2:*** *A full list of search strings used on different databases*.

***Supplementary Material 3:*** *MMAT Quality Analysis Table*.

## Notes

### Competing Interest Statement

The authors have declared no competing interest.

### Funding Statement

This study did not receive any external funding.

## References

1. Damianidou E, Mouratidou L, Kyrousi C. Research models of neurodevelopmental disorders: The right model in the right place. Frontiers in Neuroscience. 2022;16:1031075.

2. Hadders-Algra M. Early Diagnostics and Early Intervention in Neurodevelopmental Disorders---Age-Dependent Challenges and Opportunities. Journal of Clinical Medicine. 2021;10(861).

3. Frances L. Current state of knowledge on the prevalence of neurodevelopmental disorders in childhood according to the DSM-5: A systematic review in accordance with the Prisma Criteria. Child and Adolescent Psychiatry and Mental Health. 2022;16(1).

4. Association AP. Diagnostic and Statistical Manual of Mental Disorders: DSM-5. Arlington, VA; Washington, DC: American Psychiatric Association; 2013.

5. Snowling MJ, Hulme C. Annual research review: the nature and classification of reading disorders--a commentary on proposals for DSM-5. Journal of Child Psychology and Psychiatry. 2012;53(5):593–607.

6. Castaldi E, Piazza M, Iuculano T. Learning disabilities: Developmental dyscalculia. Handbook of Clinical Neurology. 2020;174:61–75.

7. Chung PJ, Patel DR, Nizami I. Disorder of written expression and dysgraphia: definition, diagnosis, and management. Translational Pediatrics. 2020;9(Suppl 1):S46–S54.

8. Braithwaite EK, Gui A, Jones EJH. Social attention: What is it, how can we measure it, and what can it tell us about autism and ADHD? Progress in Brain Research. 2020;254:271–303.

9. Jepsen P, Younossi ZM. The global burden of cirrhosis: A review of disability-adjusted life-years lost and unmet needs. Journal of Hepatology. 2021;75(Suppl 1):S3–S13.

10. FragaGonzlez G, Karipidis II, Tijms J. Dyslexia as a Neurodevelopmental Disorder and What Makes It Different from a Chess Disorder. Brain Sciences. 2018;8(10):189.

11. Hen-Herbst L, Rosenblum S. Handwriting and Motor-Related Daily Performance among Adolescents with Dysgraphia and Their Impact on Physical Health-Related Quality of Life. Children. 2022;9(10):1437.

12. Reddy SM, Reddy KJ. A comparative study of the knowledge, attitude and quality of life about dyscalculia among adolescents with and without dyscalculia. Journal of Contemporary Psychological Research. 2016;4(1):36.

13. Aro T, Eklund K, Eloranta AK, Nrhi V, Korhonen E, Ahonen T. Associations Between Childhood Learning Disabilities and Adult-Age Mental Health Problems, Lack of Education, and Unemployment. Journal of Learning Disabilities. 2019;52(1):71–83.

14. Baig MM, GholamHosseini H, Moqeem AA, Mirza F, Lindén M. Clinical decision support systems in hospital care using ubiquitous devices: Current issues and challenges. Health Informatics J. 2019;25(3):1091–104.

15. Sahoo MK, Biswas H, Padhy SK. Psychological Co-morbidity in Children with Specific Learning Disorders. Journal of Family Medicine and Primary Care. 2015;4(1):21–5.

16. Kularatna S, Jadambaa A, Senanayake S, Brain D, Hawker N, Kasparian NA, et al. The Cost of Neurodevelopmental Disability: Scoping Review of Economic Evaluation Methods. Clinicoeconomics and Outcomes Research. 2022;14:665–82.

17. Karande S, D’Souza S, Gogtay N, Shiledar M, Sholapurwala R. Economic burden of specific learning disability: A prevalence-based cost of illness study of its direct, indirect, and intangible costs. Journal of Postgraduate Medicine. 2019;65(3):152–9.

18. Arora S, Goodall S, Viney R, Einfeld S. Societal cost of childhood intellectual disability in Australia. Journal of Intellectual Disability Research. 2020;64(7):524–37.

19. NICE. NICE Guideline NG87 -- Autism spectrum disorder in under 19s: support and management 2019 [Available from: https://www.nice.org.uk/guidance/ng87.

20. Mather N, Schneider D. The use of cognitive tests in the assessment of dyslexia. Journal of Intelligence. 2023;11(5):79.

21. Roberts G, Price A, Oberklaid F. Paediatrician’s role in caring for children with learning difficulties. Journal of Paediatrics and Child Health. 2012;48(12):1086–90.

22. Sonuga-Barke EJ, Koerting J, Smith E, McCann DC, Thompson M. Early detection and intervention for attention-deficit/hyperactivity disorder. Expert Review of Neurotherapeutics. 2011;11(4):557–63.

23. Nazarova VA, Sokolov AV, Chubarev VN, Tarasov VV, Schith HB. Treatment of ADHD: Drugs, psychological therapies, devices, complementary and alternative methods as well as the trends in clinical trials. Frontiers in Pharmacology. 2022;13:1066988.

24. Verwimp C, Vaessen A, Snellings P, Wiers RW, Tijms J. The COVID generation: Online dyslexia treatment equally effective as face-to-face treatment in a Dutch sample. Annals of Dyslexia. 2024.

25. Khaleghi A, Aghaei Z, Behnamghader M. Developing two game-based interventions for dyslexia therapeutic interventions using gamification and serious games approaches entertainment computing journal. Entertainment Computing. 2022;42:100482.

26. Bucci MP. Visual training could be useful for improving reading capabilities in dyslexia. Applied Neuropsychology: Child. 2021;10(3):199–208.

27. Stokes JD, Rizzo A, Geng JJ, Schweitzer JB. Measuring Attentional Distraction in Children With ADHD Using Virtual Reality Technology With Eye-Tracking. Frontiers in Virtual Reality. 2022;3:855895.

28. Lee TL, Yeung MK, Sze SL, Chan AS. Eye-tracking training improves inhibitory control in children with attention-deficit/hyperactivity disorder. Brain sciences. 2021;11(3):314.

29. Vajs I, Papi. Accessible Dyslexia Detection with Real-Time Reading Feedback through Robust Interpretable Eye-Tracking Features. Brain Sciences. 2023;13(3):405.

30. Jafarlou F, Jarollahi F, Ahadi M, Sadeghi-Firoozabadi V, Haghani H. Oculomotor rehabilitation in children with dyslexia. Medical journal of the Islamic Republic of Iran. 2017;31:125.

31. Takahashi N, Ono T, Omori Y, Iizumi M, Kato H, Kasuno S, et al. Assessment of executive functions using a 3D-video game in children and adolescents with ADHD. Frontiers in Psychiatry. 2024;15:1407703.

32. Fernández-Martín P, Rodríguez-Herrera R, Cánovas R, Díaz-Orueta U, Martínez de Salazar A, Flores P. Data-driven profiles of attention-deficit/hyperactivity disorder using objective and ecological measures of attention, distractibility, and hyperactivity. European child & adolescent psychiatry. 2024;33(5):1451–63.

33. Yoo JH, Kang C, Lim JS, Wang B, Choi C-H, Hwang H, et al. Development of an innovative approach using portable eye tracking to assist ADHD screening: a machine learning study. Frontiers in Psychiatry. 2024;15:1337595.

34. Gomolka Z, Zeslawska E, Czuba B, Kondratenko Y. Diagnosing Dyslexia in Early School-Aged Children Using the LSTM Network and Eye Tracking Technology. Applied Sciences. 2024;14(17):8004.

35. Oh S, Joung Y-S, Chung T-M, Lee J, Seok BJ, Kim N, et al. Diagnosis of ADHD using virtual reality and artificial intelligence: an exploratory study of clinical applications. Frontiers in Psychiatry. 2024;15:1383547.

36. Toki EI, Tsoulos IG, Santamato V, Pange J. Machine learning for predicting neurodevelopmental disorders in children. Applied Sciences. 2024;14(2):837.

37. Yepes-Nuez J, Urrutia G, Romero-Garcia M, Alonso-Fernandez S. The PRISMA 2020 statement: an updated guideline for reporting systematic reviews. Revista Espa \∼ n ola de Cardiolog \’i a (English Edition). 2021;74:790–9.

38. Hong QN, Sergi F, Gillian B, Felicity B. The mixed methods appraisal tool (MMAT) version 2018 for information professionals and researchers. Education and Information Technologies. 2018;34:285–91.

39. Booth A, Sutton A, Clowes M, Martyn-St James M. Systematic approaches to a successful literature review.

40. Apiquian R, Ulloa RE, Victoria G. Standardization and validity of Chefmania, a video game designed as a cognitive screening test for children. Humanities and Social Sciences Communications. 2020;7:51.

41. García-Baos A, D’Amelio T, Oliveira I, Collins P, Echevarria C, Zapata LP, et al. Novel Interactive Eye-Tracking Game for Training Attention in Children With Attention-Deficit/Hyperactivity Disorder. Prim Care Companion CNS Disord. 2019;21(4).

42. Garcia-Redondo P, Garca T, Areces D, Nez JC, Rodrguez C. Serious Games and Their Effect Improving Attention in Students with Learning Disabilities. International Journal of Environmental Research and Public Health. 2019;16(14):2480.

43. Garcia-Zapirain B, de la Torre Díez I, López-Coronado M. Dual system for enhancing cognitive abilities of children with ADHD using leap motion and eye-tracking technologies. Journal of medical systems. 2017;41:1–8.

44. Heller MD, Roots K, Srivastava S, Schumann J, Srivastava J, Hale TS. A Machine Learning-Based Analysis of Game Data for Attention Deficit Hyperactivity Disorder Assessment. Games for Health Journal. 2013;2(5):291–8.

45. Slobodin O, Yahav I, Berger I. A Machine-Based Prediction Model of ADHD Using CPT Data. Frontiers in Human Neuroscience. 2020;14:560021.

46. Türkan BN, Amado S, Ercan ES, Perçinel I. Comparison of change detection performance and visual search patterns among children with/without ADHD: Evidence from eye movements. Research in developmental disabilities. 2016;49:205–15.

47. Sweere DJ, Pel JJ, Kooiker MJ, van Dijk JP, van Gemert EJ, Hurks PP, et al. Clinical utility of eye tracking in assessing distractibility in children with neurological disorders or ADHD: A cross-sectional study. Brain Sciences. 2022;12(10):1369.

48. Perochon S. A tablet-based game for the assessment of Visual Motor Skills in Autistic Children. npj Digital Medicine. 2023;6(1).

49. Van Viersen S, Slot EM, Kroesbergen EH, Van’t Noordende JE, Leseman PP. The added value of eye-tracking in diagnosing dyscalculia: A case study. Frontiers in psychology. 2013;4:679.

50. Piazzalunga C, Dui LG, Termine C, Bortolozzo M, Matteucci M, Ferrante S. Investigating Visual Perception Impairments through Serious Games and Eye Tracking to Anticipate Handwriting Difficulties. Sensors. 2023;23(4):1765.

51. Alt M, Hogan T, Green S, Gray S, Cabbage K, Cowan N. Word Learning Deficits in Children With Dyslexia. Journal of Speech, Language, and Hearing Research. 2017;60(4):1012–28.

52. Ikermane M, El Mouatasim A. Dyslexia deep clustering using webcam-based eye tracking. IAES International Journal of Artificial Intelligence (IJ-AI). 2023;12:1892.

53. Jafarlou F, Ahadi M, Jarollahi F. Eye movement patterns in Iranian dyslexic children compared to non-dyslexic children. Auris Nasus Larynx. 2021;48(4):594–600.

54. Korneev A, Matveeva EY, Akhutina T. What we can learn about reading development from the analysis of eye movements. Human Physiology. 2018;44:183–90.

55. Larco A, Carrillo J, Chicaiza N, Yanez C, Lujn-Mora S. Moving beyond Limitations: Designing the Helpdys App for Children with Dyslexia in Rural Areas. Sustainability. 2021;13:7081.

56. Masulli F, Galluccio M, Gerard C-L, Peyre H, Rovetta S, Bucci MP. Effect of different font sizes and of spaces between words on eye movement performance: An eye tracker study in dyslexic and non-dyslexic children. Vision research. 2018;153:24–9.

57. Moiroud L, Gerard CL, Peyre H, Bucci MP. Developmental Eye Movement test and dyslexic children: A pilot study with eye movement recordings. PLoS One. 2018;13(9):e0200907.

58. Nilsson Benfatto M, Öqvist Seimyr G, Ygge J, Pansell T, Rydberg A, Jacobson C. Screening for dyslexia using eye tracking during reading. PloS one. 2016;11(12):e0165508.

59. Prabha AJ, Bhargavi R. Predictive model for dyslexia from fixations and saccadic eye movement events. Computer Methods and Programs in Biomedicine. 2020;195:105538.

60. Jothi Prabha A, Bhargavi R. Prediction of dyslexia from eye movements using machine learning. IETE Journal of Research. 2022;68(2):814–23.

61. Rauschenberger M, Baeza-Yates R, Rello L. A universal screening tool for dyslexia by a web-game and machine learning. Frontiers in Computer Science. 2022;3.

62. Rello L, Baeza-Yates R, Ali A, Bigham JP, Serra M. Predicting risk of dyslexia with an online gamified test. Plos One. 2020;15(12):e0241687.

63. Vajs I, Ković V, Papić T, Savić AM, Janković MM. Spatiotemporal eye-tracking feature set for improved recognition of dyslexic reading patterns in children. Sensors. 2022;22(13):4900.

64. van de Ven M. Early reading intervention by means of a multicomponent reading game. Journal of Computer Assisted Learning. 2017;33(4):320--33.

65. Bonifacci P, Tobia V, Sansavini A, Guarini A. Eye-movements in a text reading task: a comparison of preterm children, children with dyslexia and typical readers. Brain Sciences. 2023;13(3):425.

66. Schmitt KL. Learning through play: The impact of web-based games on early literacy development. Computers in Human Behavior. 2018;81:378--89.

67. Peters JL, Bavin EL, Crewther SG. Eye movements during RAN as an operationalization of the RAN-reading “microcosm”. Frontiers in Human Neuroscience. 2020;14:67.

68. Niemela. Game learning analytics for understanding reading skills in transparent writing system. British Journal of Educational Technology. 2020;51(6):2376--90.

69. UCL. Learning disabilities affect up to 10 per cent of children 2013 [Available from: https://www.ucl.ac.uk/news/2013/apr/learning-disabilities-affect-10-cent-children.

70. NHS. Attention deficit hyperactivity disorder (ADHD) 2021 [Available from: https://www.nhs.uk/conditions/attention-deficit-hyperactivity-disorder-adhd/.

71. Forbes. ADHD statistics and facts in 2024. 2023.

72. Shultz JJ. Is Dyslexia Hereditary? Family Education Network. 2008.

73. Reis A. Reading and reading-related skills in adults with dyslexia from different orthographic systems: A review and meta-analysis. Annals of Dyslexia. 2020;70(3):339--68.

74. Bishop CM. Pattern recognition and machine learning: Springer; 2016.

75. NICHD. What are reading disorders? 2020 [Available from: https://www.nichd.nih.gov/health/topics/reading/conditioninfo/disorders#:~:text=Reading%20disorders%20occur%20when%20a.

76. Snowling MJ, Hulme C. Annual Research Review: The nature and classification of reading disorders–a commentary on proposals for DSM-5. Journal of child psychology and psychiatry. 2012;53(5):593–607.

77. Milano N, Simeoli R, Rega A, Marocco D. A deep learning latent variable model to identify children with autism through motor abnormalities. Frontiers in Psychology. 2023;14:1194760.

78. Mbunge E. Sensors and healthcare 5.0: Transformative SHIFT IN virtual care through Emerging Digital Health Technologies. Global Health Journal. 2021;5(4):169--77.

79. NHS. NHS Long Term Plan 2019 [Available from: https://www.longtermplan.nhs.uk/.

