## Supplementary Material 2 for "Serious Gaming and Eye-Tracking for the Screening, Monitoring, Diagnosis and Treatment of Neurodevelopmental Disorders in Children: A Systematic Literature Review"

SCOPUS:

( TITLE-ABS-KEY ( ( "Dyscalculia" OR "dyslexia" OR "reading disorder" OR "learning disorder" OR "dysgraphia" OR "ADHD" OR "neurodevelopmental disorder" OR "executive functions" OR "writing skills" OR "reading skills" ) ) AND TITLE-ABS-KEY ( ( "videogame" OR "machine learning" OR "ML" OR "AI" OR "artificial intelligence" OR "Eye" OR "Video games" OR "Educational games" OR "Machine learning" OR "Vocal analysis" OR "Deep learning" OR "Reinforcement learning" OR "Bayesian estimation" OR "Applied gaming" OR "Clustering" OR "Serious gaming" ) ) AND TITLE-ABS-KEY ( ( "child" OR "children" OR "school" OR "infant" OR "pediatric" OR "preschool" ) ) AND TITLE-ABS-KEY ( ( "screening" OR "monitoring" OR "diagnosis" OR "clinical decision support" OR "assessment" OR "epidemiology" OR "test" OR "testing" OR "therapy" ) ) )

PUBMED:

(((("Dyscalculia"[MeSH] OR "Dyslexia"[MeSH] OR "Learning Disabilities"[MeSH] OR "Agraphia"[MeSH] OR "Attention Deficit Disorder with Hyperactivity"[MeSH] OR "Neurodevelopmental Disorders"[MeSH] OR "Executive Function"[MeSH] OR "writing skills"[Title/Abstract] OR "reading skills"[Title/Abstract]))

AND (("videogame"[Title/Abstract] OR "Video Games"[MeSH] OR "machine learning"[Title/Abstract] OR "ML"[Title/Abstract] OR "AI"[Title/Abstract] OR "artificial intelligence"[Title/Abstract] OR "Eye"[Title/Abstract] OR "Educational games"[Title/Abstract] OR "Machine learning"[Title/Abstract] OR "Vocal analysis"[Title/Abstract] OR "Deep learning"[Title/Abstract] OR "Reinforcement learning"[Title/Abstract] OR "Bayesian estimation"[Title/Abstract] OR "Applied gaming"[Title/Abstract] OR "Clustering"[Title/Abstract] OR "Serious gaming"[Title/Abstract])))

AND (("child"[Title/Abstract] OR "children"[Title/Abstract] OR "school"[Title/Abstract] OR "infant"[Title/Abstract] OR "pediatric"[Title/Abstract] OR "preschool"[Title/Abstract])))

AND (("screening"[Title/Abstract] OR "monitoring"[Title/Abstract] OR "diagnosis"[Title/Abstract] OR "Decision Support Systems, Clinical"[MeSH] OR "assessment"[Title/Abstract] OR "epidemiology"[Title/Abstract] OR "test"[Title/Abstract] OR "testing"[Title/Abstract] OR "therapy"[Title/Abstract]))))
